# Replication assessment of *NUS1* variants in Parkinson’s Disease

**DOI:** 10.1101/2020.07.13.20153122

**Authors:** Bernabe I. Bustos, Sara Bandres-Ciga, J. Raphael Gibbs, Dimitri Krainc, Niccolo E. Mencacci, Ziv Gan-Or, Steven J. Lubbe, for the International Parkinson’s Disease Genomics Consortium (IPDGC).

**Affiliations:** Ken and Ruth Davee Department of Neurology and Simpson Querrey Center for Neurogenetics, Northwestern University Feinberg School of Medicine, Chicago, Illinois, USA; Molecular Genetics Section, Laboratory of Neurogenetics, National Institute on Aging, National Institutes of Health, Bethesda, MD 20892, USA; Computational Biology Group, Laboratory of Neurogenetics, National Institute on Aging, National Institutes of Health, Bethesda, MD 20892, USA; Department of Human Genetics, McGill University, Montréal, QC, H3A 1A1, Canada; Montreal Neurological Institute, McGill University, Montréal, QC, H3A 1A1, Canada; Department of Neurology and Neurosurgery, McGill University, Montréal, QC, H3A 1A1, Canada

## Abstract

The *NUS1* gene was recently associated with Parkinson’s disease (PD) in the Chinese population. Here, as part of the International Parkinson’s Disease Genomics Consortium (IPDGC), we have leveraged large-scale PD case-control cohorts to comprehensively assess the *NUS1* association in individuals of European descent. Burden analysis of rare nonsynonymous damaging variants across case-control individuals from whole-exome and -genome datasets did not find evidence of *NUS1* association with PD. Overall, single variant tests for rare (MAF<0.01) and common (MAF>0.01) variants, including 15 PD-GWAS cohorts and summary statistics from the largest PD GWAS meta-analysis to date, also did not uncover any associations. Our results indicate a lack of evidence for a role of rare damaging nonsynonymous *NUS1* variants in PD in unrelated case-control cohorts of European descent, suggesting that the previously observed association could be driven by extremely rare population-specific variants.

## Introduction

The *NUS1* gene (human chromosome 6) encodes a dehydrodolichyl diphosphate synthase subunit. It is mainly located in the endoplasmic reticulum and functions as a regulator of protein glycosylation and promotes trafficking of LDL-derived cholesterol^1,2^. Coding *NUS1* variants have been recently associated with Parkinson’s Disease (PD) in the Chinese population^3^. The authors identified a significant burden of rare (MAF<0.01) damaging nonsynonymous variants in two large independent case-control cohorts comprising 5,098 PD cases and 4,423 controls. A replication analysis of *NUS1* variants was later performed by a second group on Chinese individuals^4^, including 494 PD patients and 478 healthy controls, and found no association with the disease. Here, as part of the International Parkinson’s Disease Genomics Consortium (IPDGC), we assessed a compendium of large-scale next generation sequencing and imputed genome-wide association PD case-control cohorts in order to investigate the contribution of damaging *NUS1* variants to PD in populations of European descent.

## Results

We first assessed the burden of rare nonsynonymous *NUS1* variants in the IPDGC whole-exome sequencing data (WES)^5^ and in the Accelerating Medicines Partnership - Parkinson’s disease initiative (AMP-PD) whole-genome sequencing data (WGS, https://amp-pd.org/whole-genome-data). After quality control, variant annotation and deleteriousness prediction, we identified two heterozygous rare variants (MAF<0.01), predicted to be damaging (CADD>12.37, meaning among the 1-10 % predicted most pathogenic variants): p.P169R and p.K214E (**Table 1**). No other protein altering variants were observed. Seven out of the 18 variants identified in the original publication were located in exon 1. The variants detected in the cohorts used here are located in exons 2 and 3. None of the risk or damaging alleles from the originally reported 18 variants were observed. To assess whether we may have missed variants in exon 1, we carried out a depth of coverage analysis across all *NUS1* exons in both datasets. Although we observed a lower average read depth in exon 1 compared to the remaining exons in the IPDGC-WES dataset, no difference was detected in the AMP-PD WGS cohort (**Supplementary Table 1**) suggesting no bias in variant identification. We identified single heterozygous carriers of variants p.P169R and p.K214E in PD cases and no carriers in controls in the IPDGC-WES data (1/1,040 cases *v*. 0/452 controls). In the AMP-PD WGS data we observed five PD cases and five controls harboring the p.P169R variant (5/1,647 cases *v*. 5/1,050 controls), and one case and one control carrying the p.K214E variant (1/1,647 *v*. 1/1,050 controls). Allele-based burden assessment for both cohorts did not reveal a significant burden of damaging *NUS1* nonsynonymous variants (IPDGC-WES, P=1.0; AMP-PD, P=0.41). The combined analysis of both datasets also showed no significant burden (P=0.58). A SKAT-O^6^ test was run as an extended burden analysis on the aforementioned cohorts, selecting rare and low-frequency (MAF<0.05) coding variants regardless of their functional prediction, and again observed non-significant results (**Supplementary Table 2**). Using high quality imputed genome-wide genotypes from IPDGC-GWAS cohorts^7^, consisting of 14,671 PD cases and 17,667 controls from 15 independent studies, we did not find any of the 18 variants from the original publication and no additional coding variants with MAF<0.01, therefore no burden analysis was performed.

**Table 1.**
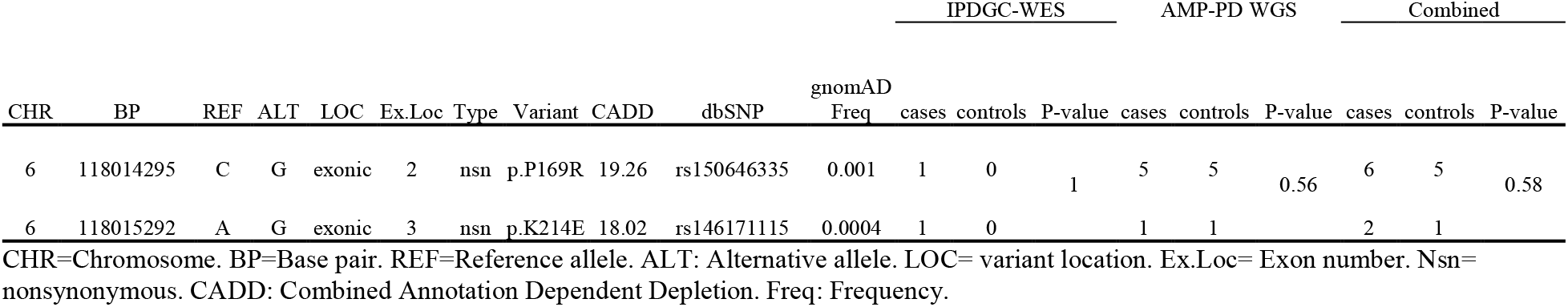
Burden analysis of rare damaging nonsynonymous variants within the *NUS1* gene in the IPDGC-WES and AMP-PD WGS datasets.

We next explored single variant associations, using rare and common variants separately within *NUS1* plus 10kb flanking regions. For rare variants we used allelic, genotypic, trend, dominant and recessive models via Fisher’s exact tests using PLINK 1.9^8^. A total of 6 rare variants were extracted from the IPDGC-WES data, of which none of them were significantly associated with the disease (P<0.05). A total of 328 rare variants were extracted from the AMP-PD WGS data, of which none survived multiple testing correction (136 linkage disequilibrium (LD)-independent variants at r^2^<0.2, Bonferroni threshold; 0.05/136 P=3.6×10^− 4^). In the IPDGC-GWAS cohorts we found 36 rare variants, of which 2 intronic variants survived multiple testing correction (rs144827068: trend P=1.62×10^−3^, allelic, dominant and genotypic P=2.4×10^−3^; rs187218668: trend P=1.73×10^−3^, allelic, dominant and genotypic P=1.8×10^−3^; 15 LD-independent variants at r^2^<0.2, Bonferroni threshold; 0.05/15 P=3.3×10^−3^). No common variants were obtained from the IPDGC-WES cohort. In the AMP-PD WGS data, we obtained 159 common variants and performed a logistic regression against disease status. We observed 2 intronic variants surpassing Bonferroni correction for 32 independent tests (rs74498762: P=1.07×10^−3^, OR=0.40, SE=0.28; rs143797072: P=1.51×10^−3^, OR=0.63, SE=0.15; Bonferroni threshold; 0.05/32 P=1.56×10^−3^) (**Supplementary Figure 1, upper panel**). In the IPDGC-GWAS cohorts, we obtained 334 common variants with none surpassing multiple testing correction (31 LD-independent at r^2^<0.2, Bonferroni threshold; 0.05/31, P=1.61×10^−3^) (**Supplementary Figure 1, middle panel**). The variants rs74498762 and rs143797072 were not observed in this dataset.

We next extracted the summary statistics for all *NUS1* variants (plus 10kb flanking regions) from the most recent PD GWAS meta-analysis^7^ (excluding 23andMe samples) that includes 15,056 PD cases, 18,618 UK Biobank proxy-cases (i.e., subjects with a first degree relative with PD) and 449,056 controls. This represents a meta-analysis of the 15 cohorts used in this study with additional cohorts. None of the 18 variants reported in the original publication were observed. We extracted 308 variants, where 15 showed uncorrected P<0.05 (**Supplementary Figure 1, lower panel**). None of the abovementioned significant variants observed in the IPDGC-GWAS cohort and AMP-PD showed association with PD risk (rs144827068, P=0.62; rs187218668, P=0.44; rs74498762, P=0.95; rs143797072, P=0.66).

We then explored the observed allele frequencies of the 18 variants from the original study in gnomAD v2.1.1^9^, encompassing 125,748 exomes and 15,708 genomes (https://gnomad.broadinstitute.org). Twelve variants do not have any reported frequencies in any representative populations, and 6 are present mostly in Asian populations (**Supplementary Table 3**).

## Conclusions

Here we have performed a comprehensive association assessment of *NUS1* variants using the largest available PD genetic datasets of European descent. We did not observe any evidence of enrichment of rare damaging nonsynonymous variants in PD cases compared to controls. At the single variant level, while we observed some suggestive associations, the largest meta-analysis of PD GWAS data^7^ did not uncover any of the *NUS1* reported variants. The lack of support presented here supports the negative results recently reported in a study performed in Chinese population^4^, however we note the limitations of being statistically underpowered to prove the null association due to small sample sizes used to detect the rare cumulative effect, therefore we cannot rule out a potential role of very rare *NUS1* variants on the risk for PD among other populations. The fact that most of the variants reported in the original population have no frequency in gnomAD, indicates that these are extremely rare, mostly present in Asian populations, and likely population specific. Additional large-scale familial and case-control studies in non-European ancestry populations are needed to further evaluate the role of *NUS1* in PD etiology.

## Methods

We utilized four sets of available PD case-control cohorts: (i) The IPDGC-WES data consists of a total number of 1,494 self-reported European individuals, comprising 1,042 PD cases and 452 neurologically healthy controls (https://pdgenetics.org/resources). A more detailed description of the cohort is reported elsewhere^5^; (ii) The AMP-PD WGS data included 1,647 PD cases and 1,050 healthy controls of European descent from three different cohorts (Biofind; https://biofind.loni.usc.edu/), Parkinson’s Disease Biomarker Program (PDBP; https://pdbp.ninds.nih.gov/), and Parkinson’s Progression Markers Initiative (PPMI; https://www.ppmi-info.org/). Cohort characteristics and quality control procedures are described in https://amp-pd.org/whole-genome-data. We have performed IBD analysis with PLINK v.1.9^8^ between the IPDGC-WES and AMP-PD data to check for overlapping individuals. We didn’t observe any duplicate samples; (iii) The IPDGC-GWAS data consisted of 15 cohorts, including a total of 32,338 individuals, comprising 14,671 PD cases and 17,667 healthy controls of European ancestry. Quality control and genotype imputation procedures were previously described^6^; and (iv) Summary statistics from the most recent PD GWAS meta-analysis^7^ excluding 23andMe data, were leveraged to explore associations with disease status. Details on quality control and imputation procedures are described in the original publication.

Variant annotation for *NUS1* was performed using ANNOVAR^10^. First, we extracted all variants within 10 kilobases up- and downstream from the start and end of the gene, respectively. Then, we selected nonsynonymous variants with MAF<0.01. Variants with CADD scores > 12.37 were selected as “damaging”^11^. Variants with MAF<0.01 in coding regions, regardless of their functional prediction, were included in the extended burden tests. All variants with MAF<0.01 and MAF>0.01 were selected separately for single variant association tests.

The burden test for rare nonsynonymous damaging variants in the IPDGC-WES and AMP-PD WGS data was assessed taking the sum of allele counts in cases and controls, separately, and built 2×2 contingency tables to perform a Fisher’s exact test. We also used SKAT-O^6^ in RVTESTS^12^ to assess variant enrichment in cases *v*. controls, adjusting for sex, population structure (PCs 1-10) and age if applicable. For single rare variant tests (MAF<0.01), we assessed allelic, genotypic, trend, dominant and recessive models with a Fisher’s exact test using PLINK. For single common variant tests (MAF>0.01) across all genotyping datasets, we used a logistic regression using PLINK, accounting for sex, age of onset in cases and age of recruitment in controls, population structure (PCA eigenvectors 1-10) and cohort if applicable. Multiple testing correction was used after calculating the total number of LD-independent variants within the gene (r^2^<0.2) and performing a Bonferroni correction to the obtained P-values. Depth of coverage assessment of all *NUS1* exons in IPDGC-WES and AMP-PD data was based on both mean depth of coverage from the aligned reads over *NUS1* exons and the mean depth of coverage per variant within the exons of *NUS1*.

## Data Availability

All datasets used in the present study are publicly available from the respective studies and databases. The IPDGC datasets including whole-exome sequencing, GWAS and summary statistics from the 2019 PD meta-analysis were obtained from https://pdgenetics.org/resources. The AMP-PD whole-genome sequencing cohort was obtained from https://amp-pd.org/.

https://github.com/ipdgc/IPDGC-Trainees/blob/master/NUS1.md

